# Corpus callosum abnormalities at term-equivalent age are associated with language development at two years corrected age in infants born very preterm

**DOI:** 10.1101/2023.09.20.23295848

**Authors:** Katsuaki Kojima, Julia E Kline, Mekibib Altaye, Beth M Kline-Fath, Nehal A Parikh, the Cincinnati Infant Neurodevelopment Early Prediction Study (CINEPS) Investigators

## Abstract

We studied the impact of microstructural abnormalities in the corpus callosum on language development in 348 infants born very prematurely. We discovered that the fractional anisotropy of the corpus callosum anterior midbody was a significant predictor of standardized language scores at two years, independent of clinical and social risk factors.

## INTRODUCTION

Preterm birth significantly increases the risk of neurodevelopmental impairments (NDIs) in children, as indicated by numerous studies ^1-3^. A particular area of concern within these impairments is language development ^4, 5^. Language outcomes carry considerable significance, as they can affect several functional outcomes ranging from academic performance and social interaction to future earning potential ^6-8^.

The early detection of NDIs is crucial for the timely initiation of appropriate interventions ^9^. However, predicting language outcomes presents a substantial challenge due to the complex nature of language development. This complexity arises from a multitude of factors including, but not limited to, socioeconomic status ^10, 11^, extent of language exposure ^12^, hearing capabilities ^13^, neurological complications, and primary speech and language disorders.

In preterm children, language outcomes have been associated with various factors, including medical complications during the neonatal period, such as bronchopulmonary dysplasia (BPD) ^14, 15^ and sepsis ^16^. Additionally, neuroimaging abnormalities discernible in brain MRIs at term ^17, 18^ as well as socioeconomic elements ^19^ are understood to influence language outcomes.

Diffusion tensor imaging (DTI) offers a method to assess the mobility of water molecules within tissues, facilitating the visualization and evaluation of white matter tract microstructure ^20^. Notably, microstructural changes in white matter, as identified by DTI, have been correlated with language outcomes in children born preterm ^21-24^. In particular, the corpus callosum (CC) has been consistently associated with language outcomes of preterm children ^22^. Fractional anisotropy (FA) values in the CC specifically have been linked to language outcomes even after accounting for medical risks ^25^. Within the CC, the anterior midbody of the CC appears to be particularly correlated with neurodevelopmental outcomes ^26^. However, the potential predictive value of DTI measures in the anterior midbody of the CC for language outcomes - independent of clinical, social, and term MRI abnormalities - has yet to be thoroughly investigated.

A significant challenge in conducting DTI studies is the manual analysis of DTI data, which can be both time-intensive and susceptible to measurement errors. An automated tractography segmentation (TractSeg) tool ^27^ was developed to address these challenges. This openly-available tool uses an encoder-decoder fully convolutional neural network (FCNN) for segmentation. The model was trained and evaluated on a dataset of 105 subjects from the Human Connectome Project (HCP), where reference segmentations for 72 tracts were obtained through a multi-step process involving existing tractography methods and manual refinement. Wasserthal and colleagues showed that the trained model demonstrated remarkable accuracy in segmenting white matter tracts, even when dealing with reduced-quality data, and outperformed a variety of other available methods. This approach offers improved reproducibility and significantly reduces the likelihood of measurement errors. The TractSeg tool has already been utilized to evaluate white matter tracts in school-age preterm children ^28^, patients with traumatic brain injuries ^29^, and gliomas ^30^.

In the present study, we aim to investigate the correlation between microstructural white matter abnormalities in the anterior midbody of the CC and language outcomes in very preterm infants using TractSeg software. Our hypothesis is that the FA values in the anterior midbody of the CC can serve as a predictor of language development at age two, independently of clinical, neuroimaging, and social risk factors.

## METHODS

The present study is a secondary analysis utilizing data gathered prospectively from the Cincinnati Infant Neurodevelopment Early Prediction Study cohort of very preterm infants.

Comprehensive details about the cohort study have been described elsewhere ^15^. In brief, over 300 infants born very preterm (at or before 32 weeks of gestational age) were recruited from 5 neonatal intensive care units (NICUs) in the greater Cincinnati area. All infants born very preterm in one of these NICUs between September 2016 and November 2019 were eligible for inclusion. We approached 95% of all eligible very preterm infants. Infants were excluded if they met any of the following criteria: (1) known chromosomal or congenital anomalies affecting the central nervous system; (2) cyanotic heart disease; or (3) hospitalization and mechanical ventilation with greater than 50% supplemental oxygen at 45 weeks postmenstrual age (PMA).

The Cincinnati Children’s Hospital institutional review board approved the study, and the review boards of the other participating hospitals approved the study based on established reciprocity agreements. Written informed consent was provided by a parent or guardian of each study infant after they were given at least 24 hours to review the consent and ask questions of the investigators.

A trained team of research personnel collected a predefined list of maternal characteristics, details related to pregnancy/delivery, and infant data ^15^. Several known predictors of neurodevelopmental outcomes were evaluated in this study, which included gestational age at birth, severe brain abnormality on MRI, post-menstrual age (PMA) at the time of MRI, sex, high-risk social status, severe BPD (Grade III) ^31^, and culture-positive late-onset sepsis.

Social status was evaluated using a composite metric consisting of six aspects: family structure (reflecting the number and relational context of the child’s caregivers), the educational level of the primary caregiver, the occupation of the main income earner, household income, language spoken at home, and maternal age at the time of birth ^32^. The scores for this metric ranged from 0 to 12, with a score of 6 or higher designated as high risk.

Brain MRIs were conducted between 39 and 44 weeks PMA. Images were collected during natural sleep with a 3T Philips Ingenia scanner (Philips Healthcare) and a 32-channel head coil located at Cincinnati Children’s Hospital, as previously described ^33^. All participants underwent their scans at this single site. DTI was obtained using a 36-direction spin echo EPI sequence with parameters set as follows: TE 88 ms, TR 6972 ms, FOV 160 × 160 mm, in-plane resolution of 2mm x 2mm, slice thickness of 2mm, and a b-value of 800 s/mm^2^. To mitigate the need for sedation, we implemented our ‘feed-and-wrap’ technique along with a fast spin echo EPI sequence with multiband imaging to minimize the time infants in the scanner and the effects of motion. We used the recently developed Developing Human Connectome Project (dHCP) pipeline^34^ for dMRI preprocessing, as previously described^35^. This pipeline reduces the effects of head motion by 1) selecting the “best” pair (i.e., least affected by intra-volume motion) of b0 volumes for each PE direction; 2) applying FSL EDDY, a non-parametric approach that corrects for distortions from motion, motion-induced signal drop-out, and eddy currents; and 3) detecting and replacing outlier slices.

We utilized a composite MRI abnormality severity score, known as the global brain abnormality score, as outlined by Kidokoro et al. ^36^ and as previously described in our cohort^37^. This score is determined by signal abnormalities and instances of impaired brain development across multiple regions, including the cerebral white matter, cortical gray matter, deep nuclear gray matter, and cerebellum^36^. All MRI scans and two-dimensional biometric measurements were conducted by a single pediatric neuroradiologist who was responsible for computing the global brain abnormality score. This neuroradiologist was blind to all clinical data to ensure objectivity in the assessments.

For each subject, we preprocessed the diffusion-weighted data as previously described ^17, 38^. Preprocessing included eddy current correction and rotation of the b-vectors. We input the diffusion images into TractSeg ^27^ with the ‘--raw_diffusion_input’ flag, which alerts TractSeg that the input is diffusion images rather than peak images (the default). TractSeg then performs constrained spherical deconvolution (CSD) to extract the three principal fiber orientations at each voxel. Next, TractSeg segments the start and end regions of the bundles to create tract orientation maps (TOMs). Finally, probabilistic tractography is performed by seeding from the TOMs. The TractSeg algorithm samples orientations from a Gaussian distribution placed on each fiber orientation distribution peak. Only streamlines starting and ending in the correct regions are retained. After tractography, we extracted the fractional anisotropy (FA) values along all streamlines for 50 tracts, including seven regions of the CC (splenium, isthmus, anterior midbody, posterior midbody, rostrum, rostral body, and genu; see Figure 1, online only) as described by Teli et al. ^39^.

**Figure 1.**
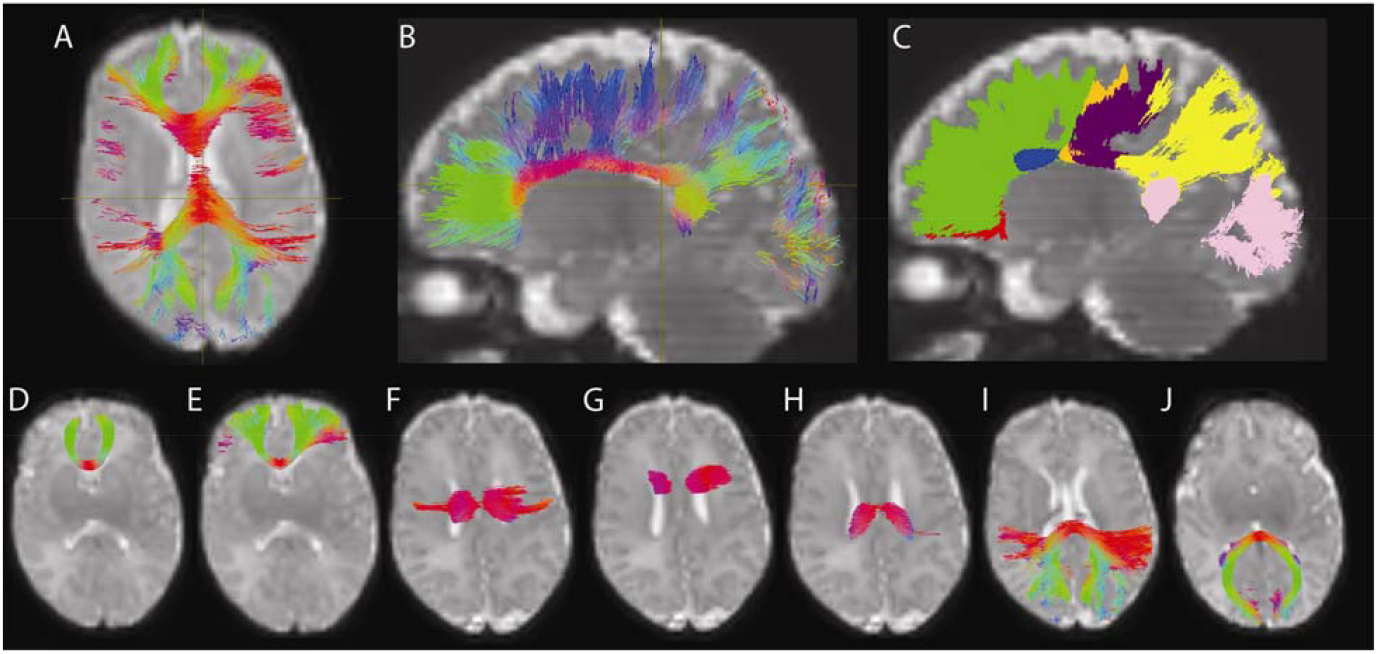
Tractography of Corpus Callosum Subregions. Axial views of all corpus callosum subregions are shown in Panel A, while sagittal views are depicted in Panel B. Panel C provides a sagittal view with color coding to differentiate each subregion: red for the rostrum, green for the genu, blue for the rostral body, orange for the anterior midbody, purple for the posterior midbody, yellow for the isthmus, and pink for the splenium. Panels D to J show the axial orientation of individual subregions: the rostrum (D), genu (E), anterior midbody (F), rostral body (G), posterior midbody (H), isthmus (I), and splenium (J), respectively.

Skilled examiners performed developmental evaluations when the children were between 22 and 26 months corrected age using the Bayley Scales of Infant & Toddler Development, 3rd edition (BSID-III) ^40^. All examiners involved in the Bayley assessments were trained in accordance with the standards of the NICHD Neonatal Research Network. The BSID-III language subscale measures language comprehension and vocabulary; the language composite score is comprised of and derived from the receptive and expressive language subdomain scores. The language composite score was normalized to have a mean of 100, a standard deviation of ±15, and a range of 40 to 160. Receptive and expressive language scores were converted into scaled scores with a mean of 10 and a standard deviation (SD) of 3 ^40^. These scores were calculated for the child’s corrected age at the assessment.

Patient demographics, FA values, and BSID-III scores were summarized using medians and ranges for continuous variables and frequencies and percentages for categorical variables. To investigate the relationship between the FA values of the anterior midbody of the CC and the composite Language score of BSID-III (primary outcome), we employed multivariable linear regression analysis, incorporating the predefined predictors of language outcome as covariates (gestational age at birth, severe brain abnormality on MRI, PMA at the time of MRI, sex, high-risk social status, severe BPD (Grade III) ^31^, and culture-positive late-onset sepsis). In our secondary analyses, we evaluated additional CC segments as predictors, with receptive and expressive language scores serving as additional outcome variables. To assess any potential bias in our findings, we conducted a comparison of baseline characteristics between the study participants who completed the study and those lost to follow-up.

## RESULTS

Out of the 348 infants enrolled in the study with identical DTI data acquisition, high-quality DTI at term-equivalent age was obtained from 328 (94%) of them. Sixteen participants could not provide this data due to poor-quality images resulting from severe brain injury or motion artifacts, and four participants were excluded - two withdrew from the study, one died prior to BSID-III testing at a year of age, and one was identified as an outlier due to extensive hemorrhagic injury. A total of 280 infants (85%) successfully completed the Bayley assessment. The baseline characteristics, FA values of the six regions of the CC, and language outcomes are presented in Tables 1 and 2 (online only).

**Table 1:** Clinical characteristics for very preterm infants with diffusion tensor imaging at term-corrected age.

|  |  |
| --- | --- |
|  | n = 328 |
| Gestation, weeks, median (range) | 29.8 (23.0, 32.9) |
| Postmenstrual age at DTI, weeks, median (range) | 43.0 (39.0, 45.1) |
| Birth weight, grams, median (range) | 1235 (410, 2905) |
| Birth weight, z-score, median (range) | 0.0 (-3.5, 3.8) |
| Race<sup>†</sup> |  |
| Black | 75 (24%) |
| White | 216 (69%) |
| Other | 23 (7%) |
| Hispanic<sup>†</sup> | 21 (7%) |
| Female sex | 159 (48%) |
| Any antenatal steroid exposure | 300 (91%) |
| Apgar scores <5 at 5 minutes<sup>‡</sup> | 44 (14%) |
| Intraventricular hemorrhage grade II, III, or IV, or any cerebellar hemorrhage on cranial ultrasound within 10 days after birth | 29 (9%) |
| Grade 3 BPD | 14 (4%) |
| Postnatal steroid exposure for BPD | 31 (9%) |
| High-risk social status | 59 (18%) |
| Primary language other than English<sup>§</sup> | 20 (6%) |
| Late onset sepsis | 36 (11%) |
| Necrotizing enterocolitis | 19 (6%) |
| Severe brain abnormalities (global brain abnormality score $\geq 12$) on term MRI | 38 (12%) |

\*All values are N (%) unless otherwise noted.
<sup>†</sup>Data missing for 14 infants.
<sup>‡</sup>Data missing for 4 infants.
<sup>§</sup>Data missing for 16 infants.

**Table 2:** Mean fractional anisotropy values of the white matter tracts and language outcomes for very preterm infants.

|  | median (range) |
| --- | --- |
| Splenum | n = 325 |
|  | 0.34 (0.12 – 0.45) |
| Isthmus | n = 326 |
|  | 0.29 (0.13 – 0.38) |
| Anterior midbody | n = 289 |
|  | 0.28 (0.18 – 0.36) |
| Posterior midbody | n = 321 |
|  | 0.28 (0.18 – 0.36) |
| Rostrum | n = 314 |
|  | 0.29 (0.13 – 0.38) |
| Rostral body | n = 251 |
|  | 0.25 (0.17 – 0.32) |
| Genu | n = 324 |
|  | 0.24 (0.10 – 0.34) |
| Bayley language composite score | n = 280 |
|  | 94 (46 – 153) |
| Bayley receptive language score | n = 272 |
|  | 9.0 (1 – 26) |
| Bayley expressive language score | n = 272 |
|  | 9.0 (1 – 19) |

The comparison of baseline characteristics between infants evaluated at follow-up (n = 280) and those lost to follow-up (n = 48) indicated that these two cohorts were generally similar (p >.05), with the exception of two variables: gestational age at birth and sex (refer to Table 3, online only). Infants evaluated at follow-up had a lower gestational age, which could potentially have a negative effect on their language scores. Conversely, a higher proportion of the infants evaluated at follow-up were female, a factor that may have a favorable influence on language outcomes.

**Table 3:**
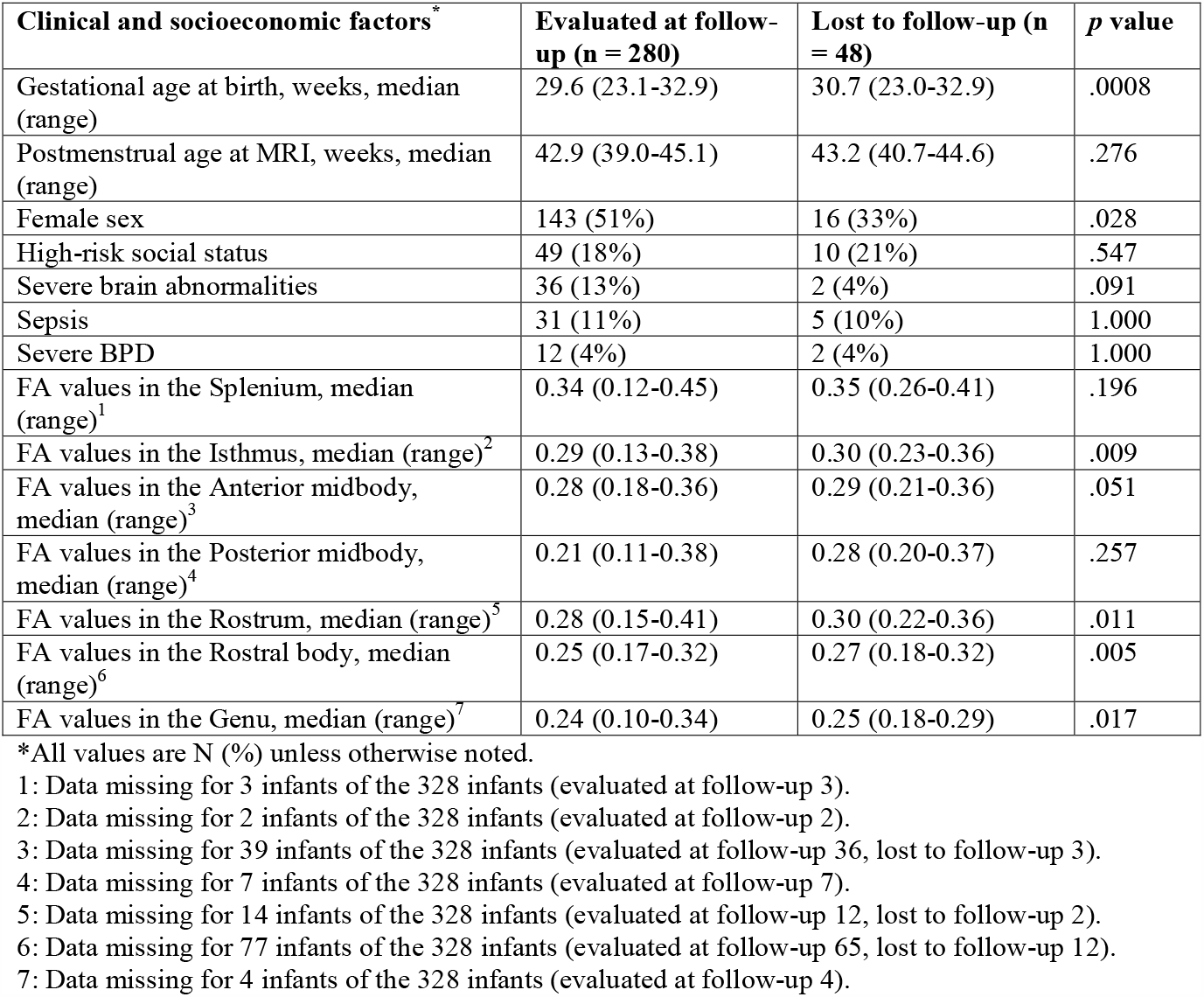
Distribution of important clinical and socioeconomic factors and DTI measures between those who were evaluated at follow-up and lost to follow-up.

The multivariable models revealed that the mean FA values in the anterior midbody of the CC were a significant predictor of Bayley language composite score (p = .047, adjusted R^2^: .182) over and above other known clinical predictors. Female sex, PMA at DTI, high-risk social status, and severe brain abnormalities at term were also significant predictors in the primary analysis for language composite scores. In secondary analyses focused on language subdomains, the FA values in the anterior midbody of the CC also emerged as a significant predictor of the Bayley expressive language score (p = .045, adjusted R^2^: .141) (Table 4), but not receptive language scores (p = .112). Additional CC segments, including the splenium (p = .048, adjusted R^2^: .158) and posterior midbody (p = .043, adjusted R^2^: .152) were found to significantly predict expressive language scores as well (Tables 5, online only).

**Table 4:** Multivariable linear regression models for predicting language development.

|  | Bayley language composite score |  | Bayley expressive language score |  |
| --- | --- | --- | --- | --- |
| Predictors | Coefficient (95% CI) | p value | Coefficient (95% CI) | p value |
| Mean FA of the anterior midbody of the corpus callosum | 96 (1, 190) | .047 | 17.1 (0.4, 33.9) | .045 |
| Female sex | 4.8 (0.2, 9.4) | .039 | 1.0 (0.2, 1.8) | .015 |
| Gestational age at birth | 0.6 (-0.4, 1.6) | .222 | -0.0 (-0.2, 0.1) | .643 |
| Postmenstrual age at DTI | -1.9 (-3.9, -0.0) | .048 | -0.3 (-0.7, 0.0) | .051 |
| High-risk social status | -14 (-20, -8) | <.0001 | -2.4 (-3.4, -1.3) | <.0001 |
| Severe brain abnormalities | -12 (-21, -2) | .015 | -1.1 (-2.8, 0.7) | .237 |
| Sepsis | -2.9 (-11.3, 5.5) | .495 | -0.5 (-2.1, 1.1) | .542 |
| Severe BPD | -0.9 (-14.8, 13.0) | .894 | -1.2 (-4.1, 1.6) | .389 |
| Adjusted R<sup>2</sup> | .182 |  | .143 |  |
| Model Statistics | F(8, 235) = 7.74, p < .0001 |  | F(8, 227) = 5.90, p < .0001 |  |

**Table 5:** Multivariable linear regression models for predicting language development.

| Predictors | Bayley expressive composite score |  | Bayley expressive language score |  |
| --- | --- | --- | --- | --- |
|  | Coefficient (95% CI) | p value | Coefficient (95% CI) | p value |
| Mean FA of the splenium of the corpus callosum | 9.5 (0.09, 19) | .048 |  |  |
| Mean FA of the posterior midbody of the corpus callosum |  |  | 14.4 (0.4, 28.4) | .043 |
| Female sex | 1.1 (0.3, 1.9) | .005 | 1.1 (0.4, 1.9) | .004 |
| Gestational age at birth | -0.1 (-0.2, 0.1) | .404 | -0.0 (-0.2, 0.1) | .588 |
| Postmenstrual age at DTI | -0.2 (-0.5, 0.1) | .164 | -0.3 (-0.6, 0.0) | .095 |
| High-risk social status | -2.4 (-3.4, -1.4) | <.0001 | -2.2 (-3.2, -1.1) | <.0001 |
| Severe brain abnormalities | -0.9 (-2.4, 0.6) | .218 | -1.0 (-2.5, 0.5) | .201 |
| Sepsis | -0.7 (-2.2, 0.7) | .305 | -0.6 (-2.0, 0.9) | .447 |
| Severe BPD | -2.1 (-4.4, 0.2) | .075 | -1.7 (-4.1, 0.7) | .162 |
| Adjusted R2 | .158 |  | .152 |  |
| Model Statistics | F(8, 260) = 7.28, p < .0001 |  | F(8, 256) = 6.91, p < .0001 |  |

## DISCUSSION

In the present study, we demonstrated that the mean FA values in the anterior midbody of the CC, measured at term-corrected age, significantly predicted both Bayley language composite scores and expressive language scores at 2 years corrected age, after controlling for known clinical and social risk factors in our cohort of very preterm infants.

Our findings bear significance in that they demonstrate a robust association between DTI measures and language outcomes, independent of both clinical and social risk factors. This association stands in contrast to previous DTI studies, which only controlled for clinical factors ^26, 41, 42^. Social factors are widely acknowledged as potent predictors of language outcomes ^10, 11^, and this was again validated in our study. A comprehensive longitudinal study tracking 224 very preterm infants over 13 years found that socioenvironmental factors correlated with language outcomes in multivariable analysis, while biological factors did not exhibit such a relationship ^19^. Another recent study incorporated a social factor in their multivariable model for language outcomes using DTI measures but did not include clinical risk factors in the same model ^25^. Our research, on the other hand, accounts for both clinical and social factors, establishing that microstructural measures remain significant independent predictors of language outcomes when all these elements are considered.

Alongside microstructural white matter abnormalities, other factors such as social risk score, female sex, and severe brain abnormalities identified on brain MRI at term-corrected age were significantly associated with language outcomes, aligning with previous research ^25, 26^. In contrast, severe BPD ^43^ and late-onset sepsis ^44^ - factors previously found to significantly predict language outcomes - did not exhibit a statistically significant association with language outcomes in our model. This discrepancy might be attributed to the relatively low occurrence of these events in our study (Of the participants who had neurodevelopmental data, 12 (4%) had severe BPD, and 31 (11%) had sepsis.), leading to insufficient statistical power to detect a significant relationship despite the relatively large sample size of our study.

Our multivariable models’ adjusted R^2^ values were relatively low (0.182 for the Bayley language composite score and between 0.143 and 0.158 for the expressive language score), though in line with previous studies ^41^. This may suggest the potential omission of important clinical predictors of language outcomes from our models, such as home language environment^45, 46^, other imaging predictors (e.g., functional MRI measures^47^ and brain morphometry^48^), and measures of hearing^49^. Additionally, accurately quantifying language development at 2 years is inherently challenging, as evidenced by the limited association between the cognitive and language outcomes at 2 years and later childhood scores in prior studies ^50, 51^. In contrast, Valavani et al. (2021) developed a machine learning model combining clinical factors and neonatal diffusion MRI measures that achieved a higher balanced accuracy of 91% for predicting language deficits at 2 years corrected age in preterm infants ^42^. He et al. ^52^ used deep learning and a multimodal MRI approach that included DTI and resting state functional MRI to achieve 88% balanced accuracy in predicting language deficits at age 2. While their primary objective was to construct a classification model for predicting language deficits—a different approach than ours—their research underscores the potential of using diffusion MRI biomarkers in conjunction with machine/deep learning techniques to enhance prediction accuracy.

We discovered significant associations between FA values in the anterior midbody of the CC and both composite and expressive language scores but not receptive language scores. This aligns with a previous study that revealed an association between the volume of the anterior portion of CC and verbal span ability, a measure of expressive language function ^53^. While further research is necessary to understand the association between specific white matter tracts and distinct language outcomes, our finding, combined with prior studies, suggests that preterm infants exhibiting abnormalities in the anterior midbody of the CC could benefit from early language intervention, with a particular emphasis on expressive language skills.

Our study has several limitations. The TractSeg software, while validated for use in school-aged children ^28^, has limited data regarding its application in newborns. Our model incorporated severe BPD and late-onset sepsis as predictors of language outcomes based on two prior large-scale studies ^14, 16^, despite other clinical risk factors, such as fetal growth restriction and necrotizing enterocolitis being suggested in other studies ^25^. We, however, consider severe BPD and sepsis to be the most rigorously studied among these clinical risk factors and aimed to avoid excessive covariates, which could lead to overfitting and collinearity. Despite these limitations, we believe our findings are significant because of the large sample size, prospectively collected data, and the evaluation of multiple known social and clinical predictors within a single model, using automated/reproducible DTI analysis.

In conclusion, within this regional cohort of very preterm infants, we found that the FA values of the anterior midbody of the CC significantly predicted language outcomes at two years corrected age, independent of various clinical and social risk factors.

## Data Availability

All data produced in the present study are available upon reasonable request to the authors.

## ACKNOWLEDGEMENT

We sincerely thank the parents of infants that participated in our study and the Cincinnati Infant Neurodevelopment Early Prediction Study (CINEPS) Investigators: Principal Investigator: Nehal A. Parikh, DO, MS. Collaborators (in alphabetical order): Armin Allahverdy, PhD, Mekibib Altaye, PhD, Anita Arnsperger, RRT, Traci Beiersdorfer, RN BSN, Kaley Bridgewater, RT(MR) CNMT, Tanya Cahill, MD, Kim Cecil, PhD, Kent Dietrich, RT, Christen Distler, BSN RNC-NIC, Juanita Dudley, RN BSN, Brianne Georg, BS, Meredith Glover, BS, Cathy Grisby, RN BSN CCRC, Lacey Haas, RT(MR) CNMT, Karen Harpster, PhD, OT/RL, Lili He, PhD, Scott K. Holland, PhD, V.S. Priyanka Illapani, MS, Kristin Kirker, CRC, Julia E. Kline, PhD, Beth M. Kline-Fath, MD, Hailong Li, PhD, Matt Lanier, RT(MR) RT(R), Stephanie L. Merhar, MD MS, Greg Muthig, BS, Brenda B. Poindexter, MD MS, David Russell, JD, Kari Tepe, BSN RNC-NIC, Leanne Tamm, PhD, Julia Thompson, RN BSN, Jean A. Tkach, PhD, Hui Wang, PhD, Jinghua Wang, PhD, Brynne Williams, RT(MR) CNMT, Kelsey Wineland, RT(MR) CNMT, Sandra Wuertz, RN BSN CCRP, Donna Wuest, AS, Weihong Yuan, PhD. We also greatly appreciate the support of our NICU fellows, nurses, and staff, and most importantly, all the study families that made this research possible.

